# Ceftazidime therapeutic drug monitoring in patients with melioidosis

**DOI:** 10.64898/2026.08.04.26359527

**Authors:** Christopher W Reilly, Simon Smith, Josh Hanson

## Abstract

**Synopsis:** *Background:* Most patients with melioidosis receive prolonged intravenous ceftazidime during the intensive phase of their antibiotic therapy. Contemporary guidelines use weight and renal function to guide dosing, but therapeutic drug monitoring (TDM) might enable further individualisation of therapy.

*Objective:* To examine the potential utility of ceftazidime TDM in the management of melioidosis.

*Methods:* We reviewed consecutive serum free ceftazidime concentrations in patients with culture-confirmed melioidosis at an Australian referral hospital. We documented the minimum inhibitory concentration (MIC) for ceftazidime of the patients’ *Burkholderia pseudomallei* isolates. We then recorded the patients’ ceftazidime dosing regimen, their serum free ceftazidime concentration and if any adverse drug reactions occurred during their treatment.

*Results:* Trough concentrations were measured in 31 patients receiving intermittent ceftazidime dosing, while random concentrations were measured in 91 patients receiving a continuous infusion. The median (range) trough concentration:MIC ratio was 37.7 (2.7-156.6) in those receiving intermittent dosing and 47.5 (8.1-181.5) in those receiving a continuous infusion. Serum ceftazidime concentrations correlated with neurotoxicity, which was documented in 5/31 (16%) receiving intermittent dosing and in 4/91 (4%) receiving a continuous infusion. Serum ceftazidime concentrations were also higher in individuals who died from their infection than in those who survived. There was no association between ceftazidime concentrations and subsequent disease recurrence.

*Conclusion:* Current dosing recommendations for the treatment of melioidosis achieve serum ceftazidime concentrations that greatly exceed the MIC of *B. pseudomallei* in this region of Australia. TDM-guided reductions in the ceftazidime dose and/or dosing frequency may mitigate the risk of ceftazidime toxicity.

## Introduction

Melioidosis is a potentially fatal opportunistic disease of socioeconomic disadvantage which is endemic to northern Australia.^1^ Antibiotic therapy is administered in two phases: an intensive phase of intravenous antibiotics for up to 8 weeks followed by an eradication phase of oral antibiotics that are taken for 3 to 6 months.^2^ In Australia, intravenous meropenem or ceftazidime are used for the intensive phase, with meropenem usually reserved for patients with neuromelioidosis or in individuals requiring ICU support.^3^ Ceftazidime is used for patients without these features and, due to its stability in elastomeric infusers, for outpatient parenteral antibiotic therapy (OPAT).^4^ The recommended dose of ceftazidime for the treatment of melioidosis is 2g 6-hourly, which is adjusted for renal function and weight.^5^ Once patients are suitable for OPAT, they transition to 24-hour continuous infusion via elastomeric infuser with a 25% dose reduction to 6g/24-hours, an approach that is associated with excellent clinical outcomes.^6^

Most Australian *Burkholderia pseudomallei* isolates have a ceftazidime MIC of ≤2 mg/L.^7^ The effective antimicrobial activity of ceftazidime occurs when the free (unbound) concentration exceeds the organism’s MIC (fT_>MIC_) for 45-100% between intermittent doses.^8^ Extrapolating from in-vitro studies of *Pseudomonas aeruginosa*, concentrations of >4 x MIC should be targeted during continuous infusions.^9^

However, over 10% of people receiving ceftazidime for the treatment of melioidosis have an adverse drug reaction (ADR) that may be dose related.^10^ Importantly, supratherapeutic ceftazidime concentrations can cause serious neurotoxicity including delirium, myoclonic jerks and even seizures.^11^ While thresholds for toxicity are incompletely defined, serum concentrations of >50 mg/L and >78 mg/L have been proposed as a guide for clinicians.^12^

The increased availability of antimicrobial TDM enables a more tailored approach to ceftazidime dosing.^13^ We therefore examined serum ceftazidime concentrations in consecutive patients with culture-confirmed melioidosis who were receiving ceftazidime by intermittent dosing and/or by 24-hour continuous infusions. We correlated their serum ceftazidime concentrations with the development of ADRs and their clinical course.

## Methods

We reviewed all patients with culture-confirmed melioidosis managed at Cairns Hospital in tropical Australia between 4^th^ May 2022 and 3^rd^ August 2025. This time period was chosen as it was associated with expanded access to TDM at the hospital. We included patients if they had a trough ceftazidime concentration measured whilst receiving intermittent dosing and/or a random ceftazidime concentration measured whilst receiving a continuous infusion. Patients receiving intermittent dosing with a concentration that was not measured immediately prior to their next ceftazidime dose were excluded. Patients receiving haemodialysis were excluded as findings in these individuals have been presented already.^14^ In patients who had multiple ceftazidime concentrations measured, we used their initial serum ceftazidime level for the analysis.

We reviewed the patients’ electronic medical record and recorded the initial ceftazidime MIC of their *B. pseudomallei* isolate, defined by ETEST (EUCAST). We also recorded the patients’ age, height and weight, and their comorbidities, clinical presentation and outcomes as described previously.^15^ Recurrent melioidosis was categorised as recrudescence (microbiologically confirmed recurrence when targeted antimicrobial therapy should have been being taken, whether it was or not) or relapse (microbiologically confirmed recurrence after the completion of prescribed eradication therapy). Comorbidity was quantified, where necessary, using the Charlson Comorbidity Index.^16^

We recorded the ceftazidime dosing regimen, which was selected at the time of the patient’s illness by the attending infectious diseases specialist with the support of two dedicated antimicrobial stewardship and OPAT pharmacists with infectious diseases expertise. We determined retrospectively whether this was consistent with current Australian guidelines.^5^ An intermittent dose of 2 g 6-hourly in patients weighing ≥40 kg with creatinine clearance >50 ml/min was deemed appropriate. Although not stipulated in current guidelines, for patients who had their intermittent ceftazidime dose or dose frequency reduced due to renal impairment and/or low body weight, we considered a continuous infusion dose reduction of 20-30% of the total 24-hour intermittent dose to be appropriate. A continuous infusion dose of 6-8 g over 24-hours for patients with neuromelioidosis was also considered appropriate.

For each measurement of serum ceftazidime, we documented the simultaneously measured serum creatinine and recorded if the serum ceftazidime concentration triggered a change to the patient’s ceftazidime dose or dosing interval. We also determined if there were any ADRs attributed to ceftazidime. Liver function test derangement was said to be present, if there was a new rise in liver enzymes to >3 times the upper limit of normal. Neurotoxicity was said to be present if it was clearly documented by the attending clinician in the medical record.

### Statistical analysis

Data were de-identified, entered into an electronic spreadsheet (Microsoft Excel) and analysed with statistical software (Stata 18.5). Groups were compared using the Wilcoxon rank-sum test.

### Ethical approval

The study was approved by the Far North Queensland Human Research Ethics Committee (HREC/15/QCH/46-977). As the data were collected retrospectively and presented in a de-identified manner, the Committee waived the requirement for informed consent.

## Results

There were 252 patients with culture confirmed melioidosis diagnosed at the hospital during the study period; serum ceftazidime concentrations were measured on 170 occasions in 115 (46%) of these individuals. Overall, 3/115 (3%) died, while 8/115 (7%) had disease recurrence (4 had recrudescence and 4 had relapse).

### Individuals receiving intermittent dosing

A ceftazidime trough concentration was measured on 41 occasions in 31 patients receiving intermittent ceftazidime dosing. Among these 31 patients, 19 (61%) were determined retrospectively to have been prescribed a ceftazidime dose consistent with current guidelines, 10 (32%) were prescribed a lower ceftazidime dose than is recommended in current guidelines and 2 (7%) were prescribed a ceftazidime dose that exceeded the dose recommended in current guidelines.

The median (range) ceftazidime MIC of the *B. pseudomallei* isolates in these 31 patients was 1 mg/L (1-4). The median (range) initial ceftazidime trough concentration was 41 mg/L (4.2-156.6), equating to a median (range) trough concentration:MIC ratio of 37.7 (2.7-156.6) (Table 1).

**Table 1.** Clinical characteristics and results of ceftazidime therapeutic drug monitoring in patients receiving treatment for melioidosis.

|  | Intermittent dosing<br>(n=31) | Continuous<br>infusion (n=91) |
| --- | --- | --- |
| Clinical characteristics |  |  |
| Age (years) (median, IQR) | 62 (51-72) | 55 (46-67) |
| BMI (kg/m<sup>2</sup>) (median, IQR) | 22.3 (18.9-28.4) | 28.1 (22.8-32.3) |
| eGFR (mL/min/1.73 m<sup>2</sup>) (median, IQR) | 64 (26-90) | 90 (73-90) |
| CrCl <sup>a</sup> (mL/min) (median, IQR) | 56 (25-90) | 90 (63-90) |
| MIC (mg/L) of *B. pseudomallei* isolate |  |  |
| 0.5 | 0 | 1 (1%) |
| 1 | 26 (84%) | 80 (88%) |
| 2 | 4 (13%) | 8 (9%) |
| 4 | 1 (3%) | 2 (2%) |
| Concentration to MIC ratio (median, range) | 37.7 (2.7-156.6) | 47.5 (8.1-181.5) |
| Toxicity |  |  |
| Any | 8 (26%) | 12 (13%) |
| Neurotoxicity | 5 (16%) | 4 (4%) |
| Eosinophilia | 5 (16%) | 12 (13%) |
| Toxicity by concentration to MIC ratio (mg/L) (median, IQR) |  |  |
| No toxicity | 36.8 (10.9-45.2) | 49 (37.6-61.2) |
| Any toxicity | 78.3 (44.8-99.8) | 60.8 (41.0-87.3) |
| No neurotoxicity | 38.1 (16.5-52.1) | 49.0 (37.6-61.2) |
| Neurotoxicity | 92.4 (53.6-129.3) | 86.2 (67.8-98.9) |
| Dosage adjustments made due to ceftazidime concentration |  |  |
| Dose change | 8 (26%) | 10 (11%) |
| Dose decreased | 5 (16%) | 9 (10%) |
| Dose increased | 1 (3%) | 0 |
| Ceftazidime ceased or changed to meropenem | 2 (6%) | 1 (1%) |

BMI, body mass index; IQR, interquartile range, eGFR, estimated glomerular filtration rate; CrCl, creatinine
clearance; MIC, minimum inhibitory concentration
<sup>a</sup> Calculated using Cockcroft-Gault Equation

Toxicity occurred in 8/31 (26%), with neurotoxicity documented in 5/31 (16%) (Table 2). Of those with neurotoxicity, one patient was receiving ceftazidime at a dose consistent with current guidelines; 2 were receiving ceftazidime at a dose higher than is recommended in current guidelines and 2 were receiving ceftazidime at a dose lower than is recommended in current guidelines. The median (range) trough concentration of ceftazidime was higher in individuals with neurotoxicity than in those without neurotoxicity (92.4 mg/L (43.0-129.3) versus 38.1 (4.2-93.3), p=0.004).

**Table 2.** Concentrations and toxicities in patients receiving ceftazidime for treatment of melioidosis.

| **Intermittent dosing** |  |  |  |  |  |
| --- | --- | --- | --- | --- | --- |
| Ceftazidime trough concentration (mg/L) | Ceftazidime regimen | CrCl<sup>a</sup> (ml/min), weight (kg) | Dose consistent with guidelines | Toxicity | Amendment to dosing regimen |
| 38.8 | 1g 24-hourly | 17, 54 | No - lower | LFT derangement<sup>b</sup> | No |
| 43 | 2g 24-hourly | 16, 63 | No - lower | Neurotoxicity: confusion, disorientation | Yes - dose reduced to 1g 24-hourly |
| 50.1 | 2g 12-hourly | 25, 74 | Yes | LFT derangement<sup>c</sup> | No |
| 64.2 | 2g 6-hourly | 49, 41 | No - higher | Neurotoxicity: confusion, LFT derangement<sup>d</sup> | Yes - dose reduced to 2g 8-hourly |
| 92.4 | 2g 24-hourly | 18, 61 | No - lower | Neurotoxicity: confusion, LFT derangement<sup>c</sup> | Yes - dose withheld and subsequently transitioned to end of life care |
| 93.3 | 1g 12-hourly | 16, 45 | Yes | LFT derangement<sup>e</sup> | No |
| 101.9 | 2g 6-hourly | 49, 61 | No - higher | Neurotoxicity: confusion & myotonic jerks | Ceased - changed to meropenem |
| 156.6 | 2g 6-hourly | 66, 71 | Yes | Neurotoxicity: confusion/agitation | Yes - dose reduced to 2g 24-hourly and subsequently transitioned to end of life care |
| **Continuous infusion** |  |  |  |  |  |
| Ceftazidime random concentration (mg/L) | Dose administered over 24 hours | CrCl (ml/min), weight (kg) | Dose appropriate<sup>f</sup> | Toxicity | Amendment to dosing regimen |
| 24.6 | 6g | >90, 74 | Yes | LFT derangement<sup>b</sup> | No |
| 30.3 | 6g | >90, 49 | Yes | LFT derangement<sup>b</sup> | No |
| 40.5 | 6g | >90, 86 | Yes | LFT derangement<sup>b</sup> | No |
| 42.6 | 6g | >90, 47 | Yes | LFT derangement<sup>b</sup> | No |
| 49.6 | 6g | >90, 91 | Yes | Diarrhoea | No |
| 59.2 | 6g | 52, 79 | Yes | Rash | Yes - changed to meropenem |
| 62.4 | 6g | 32, 80 | No - higher | Neurotoxicity - confusion, dizziness, blurred vision | Yes - reduced to 4g infuser |
| 77.9 | 6g | 48, 73 | No - higher | Nausea/anorexia | No |
| 83.8 | 6g | 46, 64 | No - higher | Neurotoxicity- dysgeusia<sup>g</sup> | No |
| 88.5 | 8g<sup>h</sup> | >90, 75 | Yes | Neurotoxicity - left sided paraesthesia | Yes - reduced to 6g infuser |
| 102.3 | 6g | 41, 85 | No - higher | Neurotoxicity - headache with blurring & diplopia | Yes - reduced to 4g infuser |
| 106.2 | 6g | 51, 40 | Yes | Developed SJS/TEN<sup>i</sup> | Yes - reduced to 4g infuser |

CrCl: creatinine clearance; LFT: liver function tests; SJS: Stevens-Johnson syndrome. TEN: toxic epidermal
necrolysis
<sup>a</sup> Calculated using Cockcroft-Gault Equation
<sup>b</sup> Alanine aminotransferase >3 x upper limit of normal
<sup>c</sup> Aspartate aminotransferase >3 x upper limit of normal
<sup>d</sup> Alanine aminotransferase and aspartate aminotransferase >3 x upper limit of normal
<sup>e</sup> Gamma-glutamyl transferase >3 x upper limit of normal
<sup>f</sup> Based on current guidelines and a 20-30% dose reduction of recommended intermittent dose in 24-hours, extrapolated from guideline recommendation of 25% dose reduction if receiving 8g/24-hours in intermittent doses.
<sup>g</sup> Felt most likely to be neurological after exclusion of oral candidiasis and other local diagnoses

There were 3/31 (10%) who died during their intensive phase treatment; the median (range) trough concentration of ceftazidime was higher in individuals who died than in those who survived their infection (92.4 mg/L (66.6-156.6) versus 38.6 (4.2-101.9), p=0.01). Of the 28 who survived their intensive phase, recrudescent infection occurred in 2 (7%). There was no relationship between the ceftazidime trough concentration and later recrudescence (p=0.66), which in both cases was likely to have been explained by other factors (Tables 3 and 4).

**Table 3.**
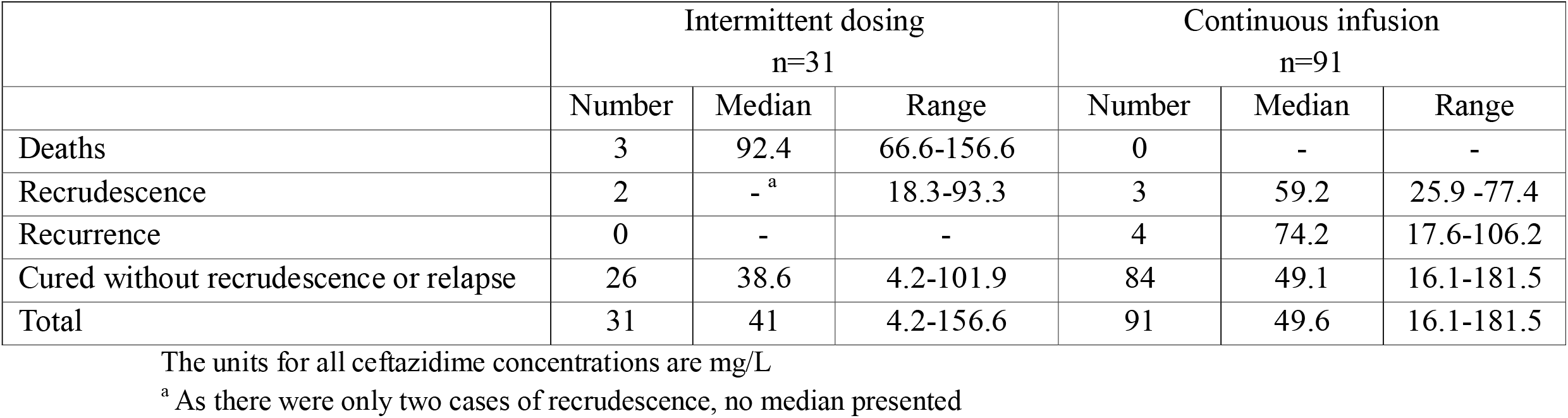
Association between results of ceftazidime therapeutic drug monitoring and the clinical course of patients receiving treatment for melioidosis.

|  | Intermittent dosing<br>n=31 |  |  | Continuous infusion<br>n=91 |  |  |
| --- | --- | --- | --- | --- | --- | --- |
|  | Number | Median | Range | Number | Median | Range |
| Deaths | 3 | 92.4 | 66.6-156.6 | 0 | - | - |
| Recrudescence | 2 | -<sup>a</sup> | 18.3-93.3 | 3 | 59.2 | 25.9 -77.4 |
| Recurrence | 0 | - | - | 4 | 74.2 | 17.6-106.2 |
| Cured without recrudescence or relapse | 26 | 38.6 | 4.2-101.9 | 84 | 49.1 | 16.1-181.5 |
| Total | 31 | 41 | 4.2-156.6 | 91 | 49.6 | 16.1-181.5 |

The units for all ceftazidime concentrations are mg/L
<sup>a</sup> As there were only two cases of recrudescence, no median presented

**Table 4.** Serum ceftazidime concentration and other factors contributing to the development of disease recrudescence and relapse patients receiving treatment for melioidosis.

| Ceftazidime regimen | Initial ceftazidime concentration (mg/L) | Ceftazidime MIC of isolate (mg/L) | Nature of recurrence | Time of culture-confirmed recurrence after completion of ceftazidime | Contributing factors |
| --- | --- | --- | --- | --- | --- |
| Intermittent <sup>a</sup> | 18.3 | 1 | Recrudescence | 82 days | Poor adherence to eradication therapy<br>Poorly controlled diabetes mellitus<br>Ongoing hazardous alcohol use |
| Intermittent | 93.3 | 1 | Recrudescence | 20 days | Doxycycline as eradication therapy <sup>b</sup><br>Poorly controlled diabetes mellitus |
| Continuous <sup>a</sup> | 77.4 | 1 | Recrudescence | 82 days | Poor adherence to eradication therapy<br>Poorly controlled diabetes mellitus<br>Ongoing hazardous alcohol use |
| Continuous | 25.9 | 1 | Recrudescence | 70 days | Doxycycline as eradication therapy <sup>b</sup><br>Osteomyelitis |
| Continuous | 59.2 | 1 | Recrudescence | 31 days | Doxycycline as eradication therapy <sup>b</sup><br>Cavitating lung abscess |
| Continuous | 97.8 | 1 | Relapse | 123 days | Empyema<br>Poor adherence to eradication therapy<br>Ongoing hazardous alcohol use |
| Continuous | 17.6 | 1 | Relapse | 143 days | Doxycycline as eradication therapy <sup>b</sup> |
| Continuous | 50.6 | 1 | Relapse | 118 days | Doxycycline as eradication therapy <sup>b</sup><br>Ongoing hazardous alcohol use |
| Continuous | 106.2 | 1 | Relapse | 160 days | Doxycycline as eradication therapy <sup>b</sup><br>Poorly controlled diabetes mellitus<br>Metastatic melanoma |

<sup>a</sup>This was the same patient
<sup>b</sup> All patients initially prescribed trimethoprim/sulfamethoxazole which was discontinued due to an adverse drug reaction

The results of the measured trough concentrations led to a change in the ceftazidime dose or dosing interval in 8/31 (26%) patients; this was a reduction in 7/31 (23%) and an increase in 1/31 (3%). Two individuals who had a dose reduction subsequently died from co-existing comorbidity after being transitioned to end-of-life care. The first, a man in his 80’s with a Charlson Comorbidity Index of 12 had a dose reduction in the setting of confusion and agitation after returning a serum ceftazidime level of 156.6 mg/L; he died from his underlying end-stage chronic obstructive pulmonary disease. The second, a man in his 90’s with a Charlson Comorbidity Index of 14, had his ceftazidime ceased in the setting of confusion, agitation and liver function test derangement after returning a serum ceftazidime level of 92.4 mg/L; he was transitioned to end-of-life care and died from multi-organ failure. There was no recrudescence or relapse in any of the individuals who had a dose reduction (Table 2).

### Individuals receiving continuous infusion

A random ceftazidime concentration was measured on 129 occasions in 91 patients receiving ceftazidime via continuous infusion. This included 7 patients who also had ceftazidime concentrations measured whilst receiving ceftazidime via intermittent dosing. Of the 91 individuals receiving ceftazidime via continuous infusion, 86 (95%) received 6g over 24-hours, 3 (3%) received 4g over 24-hours and 2 (2%) received 8g over 24-hours. On a retrospective review, which considered the patients’ weight and their creatinine clearance, it was deemed that 78 (86%) were prescribed an appropriate dose of ceftazidime, 12 (14%) had a dose greater than was appropriate and 1 (1%) had a dose that was lower than appropriate. Among the 12 individuals who had a dose that was felt to be greater than was appropriate, 9 (75%) weighed ≥60 kg and had a CrCl of 31-50 ml/min; one of the remaining three individuals had neuromelioidosis (Supplementary Table 1).

The median (range) ceftazidime MIC of the *B. pseudomallei* isolates from these 91 patients was 1 mg/L (0.5-4). The median (range) initial ceftazidime random concentration was 49.6 mg/L (16.1-181.5), equating to a median (range) random concentration:MIC ratio of 47.5 (8.1-181.5).

Toxicity occurred in 12/91 (13%) with neurotoxicity occurring in 4/91 (4%). Three of the individuals who developed neurotoxicity had a reduced creatinine clearance that would warrant an intermittent dose of 2 g 8-hourly were receiving a continuous infusion dose of 6 g over 24-hours; the other was receiving 8 g over 24-hours for the treatment of neuromelioidosis. The median (range) random concentration was higher in individuals with neurotoxicity than in those without neurotoxicity (86.2 mg/L (62.4-102.3) versus 49.0 (16.1-181.5), p=0.02). One individual receiving a 6 g continuous infusion for neuromelioidosis had a random ceftazidime level of 77.4mg/L and a paired random cerebrospinal fluid level of 9.6mg/L.

There were no deaths among the 91 patients receiving a continuous ceftazidime infusion, 3/91 (3%) had recrudescence and 4 (4%) had relapse. However, there was no relationship between the ceftazidime random concentration and disease recurrence: median (range) ceftazidime concentration (p=0.57); in all seven cases other factors (including the use of doxycycline eradication therapy, incomplete adherence, and a more complicated clinical phenotype) appeared likely to have contributed to the recurrence (Tables 1 and 4).

The random ceftazidime concentration led to a dose or dosing interval change in 10 (11%) patients; this was a dose reduction in all cases. Among the 10 individuals who had a dose reduction, 9 (90%) had no recrudescence or recurrence but one patient with metastatic melanoma had culture-confirmed recurrence of melioidosis six months after her initial presentation. The risk of relapse was increased in this case by poorly controlled immunotherapy-induced type I diabetes and the requirement to take doxycycline for her eradication therapy after she developed the Stevens-Johnson syndrome/toxic epidermal necrolysis while taking trimethoprim/sulfamethoxazole.

## Discussion

Current dosing recommendations for ceftazidime for the treatment of melioidosis achieve serum concentrations that greatly exceed the organism’s MIC in this region of Australia.^5^ In all patients receiving intermittent dosing, the fT>MIC far surpassed 100% and all patients receiving continuous infusion had a concentration >4 x MIC. While the risk of toxicity is mitigated somewhat by the wide therapeutic index of ceftazidime, the high serum concentrations seen in our study – and importantly the documentation of neurotoxicity in 7% of the patients, which was correlated with serum ceftazidime levels – suggests there is a role for TDM in defining the optimal dose of ceftazidime in patients with melioidosis.

Acknowledging that serum concentrations are higher than concentrations in deep tissues,^17^ the very high serum concentrations seen in this study suggest that adequate tissue concentration is still likely.^18^ The rate of disease recurrence was similar to previous large Australian series,^20, 21^ and was unrelated to the measured ceftazidime level. It was instead linked to more to issues with adherence, the use of doxycycline as an eradication agent and clinical phenotype including osteomyelitis and empyema which are all well recognised to increase the risk of disease recurrence.^22-24^

The single patient who had paired serum and CSF measurements had concentrations that were well above target suggesting that ceftazidime may have a role in the management of neuromelioidosis – a manifestation that occurs in 3% of melioidosis cases and where only meropenem is currently recommended.^19^ In this context, it was relevant that none of the five individuals with CNS involvement receiving ceftazidime during their intensive phase had any recrudescence of relapse of their neurological disease.

Despite the high concentrations recorded in this study, ADRs were usually mild and patients were able to be monitored with minor dosage adjustments. In only one individual with neurotoxicity was it necessary to cease ceftazidime; other patients with neurotoxicity were able to be managed with dose reduction. This observation may explain why the attending clinicians prescribed a continuous ceftazidime dose in 14% of the patients that was on review, perhaps, higher than indicated for their weight and renal function. It was also notable that 18/25 (72%) patients without any evidence of neurotoxicity had a ceftazidime concentration >78mg/L, suggesting this previously proposed threshold requires further evaluation.^12^

Ceftazidime delivered via outpatient elastomeric infusers are an established option for the treatment of melioidosis in northern Australia. However, concerns exist regarding ceftazidime’s stability and the potential for pyridine production occurring from hydrolysis in aqueous solution. Some centres recommend avoiding exposure of ceftazidime to temperatures >25°C and changing infusers every 12 hours.^25^ In our setting, ambient temperatures regularly exceed 30°C and ceftazidime elastomeric infusers are administered over a 24-hour period. The ceftazidime concentrations reported here, and the achieved outcomes support this approach, however, the high concentration/MIC ratios in this study may support a reduction in the 24-hour infusion dose if pyridine toxicity is a clinical concern.^26^

Additionally, whilst elastomeric infusers are available in Australia’s well-resourced healthcare setting, the high ceftazidime concentrations seen in those receiving intermittent dosing may have implications for OPAT in low- and middle-income countries where most melioidosis cases occur. Target attainment could possibly be achieved with reduced dosing intervals, particularly in patients with low body weight or impaired renal function, thereby facilitating hospital discharge and administration of intermittent dosing via an outpatient clinic.

Limitations of this study include its retrospective nature and the reliance on documentation from medical records. Ceftazidime trough concentrations had to be cross referenced with the timing of sample collection and dosing of ceftazidime recorded on the medication chart; any incorrect entries would give a result that was not a true trough. Random concentrations and routine blood tests are collected peripherally at our centre, not from a PICC line. However, if a patient declines peripheral phlebotomy, a PICC line sample is collected. It is therefore possible that if a PICC line sample was collected this could result in a higher ceftazidime concentration, although the standardised protocol is to discard the first 10 ml of blood that is collected in this manner. The high concentration:MIC ratios in this study can partly be explained by the low MIC of *B. pseudomallei* isolates in northern Australia which may not be generalisable to other countries. The high concentration:MIC ratios in this study do not obviate the requirement for diligent source control to achieve optimal outcomes.^24, 27^

Acknowledging these limitations, this study suggests that TDM may have utility in the treatment of melioidosis in northern Australia, enabling a more individualised approach to the use of ceftazidime. Knowledge of serum ceftazidime concentrations may help confirm a clinical suspicion of neurotoxicity, while satisfactory levels may allow reductions in both the dose of and the frequency of ceftazidime administration. This may mitigate concerns over ceftazidime related pyridine toxicity in elastomeric infusers. It may also facilitate increased accessibility to intermittent dosing administered as an outpatient in locations where elastomeric infusers are not accessible.

## Acknowledgements

The authors would like to acknowledge all the health workers involved in the care of the patients.

## Transparency declarations

### Data availability statement

Data cannot be shared publicly because of the Queensland Public Health Act 2005. Data are available from the Far North Queensland Human Research Ethics Committee (contact via) for researchers who meet the criteria for access to confidential data.

---

**Supplementary table 1.**
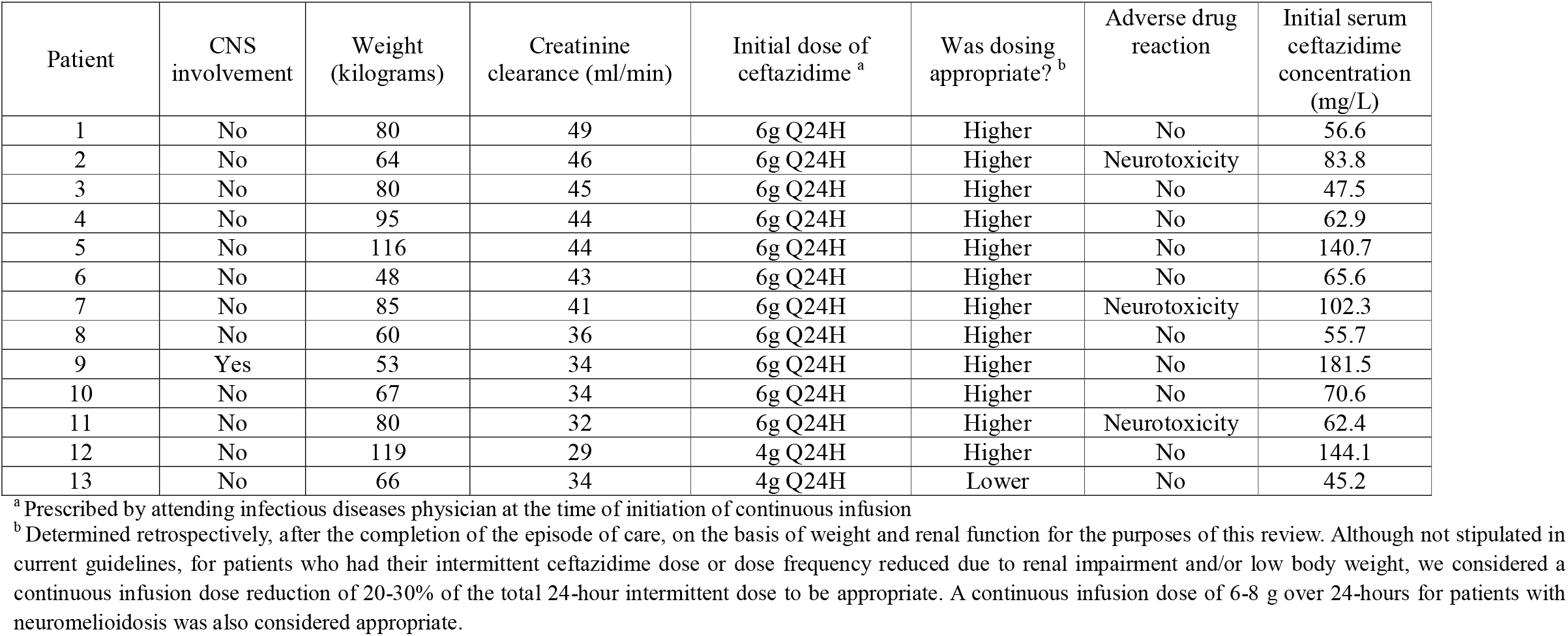
Characteristics of the individuals in whom it was determined that the dose of the continuous infusion may not have been appropriate.

